# Exploring Reproductive Violence in a Moroccan Context: A Stakeholder informed Delphi Study

**DOI:** 10.1101/2025.06.25.25330318

**Authors:** Meg Ryan, Nihal Said, Jeanne Mantel, Mina Sabih, Rania Abu Elhassan, Frédérique Vallieres, Greg Sheaf, Seri Wendoh, Abdellatif Maamri

## Abstract

**Background:** Reproductive violence (RV) is not currently recognised as a legally or epistemically distinct category of gender-based violence. While reproductive coercion (RC) as a subset of RV is gaining increased recognition within the literature, studies are predominantly focused on RC as a form of intimate partner abuse. This is despite evidence that actions taken to impede reproductive autonomy can be perpetrated at family, community, and even state level. The current study, based in Morocco, firstly aimed to examine how RV is influenced by social and cultural context. We then use this knowledge to develop a series of item statements for inclusion in a newly developed RC and RV screening tool.

**Methods:** A Delphi study was conducted with an international panel of 11 experts in the field of gender-based violence prevention and sexual and reproductive health. The study consisted of two rounds, with 90% of respondents completing all two surveys. Experts were asked to rate 33 item statements relating to forms of reproductive coercion and violence that women in Morocco may face across a five-point scale ranging from strongly agree to strongly disagree. Participants were encouraged to clarify their rating with open text responses. A 75% agreement rate was used as threshold for consensus.

**Results:** A consensus was reached for items describing the different forms of reproductive coercion and violence that women in Morocco may face. Statements largely consisted of three categories: interpersonal, institutional, and social norms.

**Conclusions:** Using contextualised screening tools for reproductive coercion and violence that consider context specific gender and social norms allows for a deeper understanding of gender-based violence in Morocco. Following further testing, this screening tool has the potential to increase the visibility of RV, inform more targeted and effective interventions, and strengthen both screening and monitoring and evaluation. The current study represents a crucial first step in the development of such a screening tool.

## Introduction

Sexual and gender-based violence (SGBV)—particularly violence against women, girls, and transgender individuals—is a widespread human rights violation with devastating effects on individuals, families, and communities (Raftery et al., 2022). With an estimated 30% of women having been subjected to physical and/or sexual violence by an intimate partner or non-partner sexual violence, or both (WHO, 2022), SGBV ranks among the leading causes of death and disability for women worldwide. Adolescents and young women face a higher risk of intimate partner violence (IPV); prevalence among ever-partnered adolescent girls aged 15–19 ranges from 16% in the last year to 24% in their lifetime (Wang et al., 2024). Women with disabilities are up to ten times more likely to experience SGBV, and individuals with diverse sexual orientations and gender expressions face significant violence and access barriers (UNFPA, 2018). In 2023, an estimated 612 million women and girls lived within 50 km of a conflict. In the same year, the UN have verified 3688 incidents of conflict-related sexual violence, with women and girl survivors making up 95% of these incidents (UN Women, 2024).

A closely related yet distinct form of SGBV is reproductive violence (RV). Defined as ‘any form of abuse, coercion, exploitation or violence that compromises reproductive autonomy and self-determination and/or violence directed at people because of their actual or perceived reproductive capacity’ (Laverty & De Vos, 2021, p.12), RV represents a significant barrier to achieving universal access to sexual and reproductive health (SRH) services. SRH needs are reflected in multiple Sustainable Development Goals (SDG) including SDG 3 (Good Health and Well-Being), through targets ensuring universal access to sexual and reproductive health services (Target 3.7), and through SDG 5’s (Gender Equality) eliminating violence against women and girls (Target 5.2) and protecting reproductive rights (Target 5.6) (UN, 2015).

A concept closely related to RV, but which has received more direct attention within the academic literature is *reproductive coercion* (RC). First coined by Miller et al., (2010), RC refers to any deliberate actions taken to influence or control the reproductive autonomy and choices of another person (Miller et al., 2010; Tarzia & Hegarty, 2021). Commonly conceptualizedj as an act perpetrated against a person by their intimate partner, forms of RC identified within the literature include contraceptive sabotage (hiding, damaging, or interfering with contraceptive use) and pregnancy coercion (pressuring someone to continue with or terminate a pregnancy) (Grace & Anderson, 2018; Silverman & Raj, 2014). Intimate partners are not the sole perpetrators of reproductive violence and coercion, however, with evidence to demonstrate that RC is also carried out by relatives including mothers-in-law, mothers, and sisters-in-law (Clark et al., 2008; Grace & Anderson, 2018; Gupta et al., 2012; McCauley et al., 2014; Muñoz et al., 2023; Park et al., 2016). Moreover, social norms can also influence societal and cultural ideas about parenting and reproductive choices, resulting in communities perpetrating RC through gender norms that legitimise control of sexuality and reproductive behavior, particularly for women (Ouahid, 2023). Finally, governments and institutions can also deliberately interfere with reproductive autonomy through upholding laws and policies that reduce access to contraception, enforce sterilization, and/or restrict access to safe abortion (Chadwick & Jace Mavuso, 2021).

While state and systemic-level restrictions on reproductive autonomy are frequently classified within the literature as forms of RV rather than RC, the distinction between these concepts remains ill-defined, with ongoing ambiguity surrounding their precise definitions (Chadwick & Mavuso 2021). Similarly, although both RV and RC constitute violations of reproductive autonomy - a right protected by international law - they are commonly viewed as distinct yet interrelated forms of SGBV as well as broader infringements on SRHR. Consequently, many legal and policy responses fail to address these specific categories as discrete forms of harm.

In Morocco, persistently high rates of GBV over the past two decades have highlighted the need for greater awareness of the need for comprehensive SRH services (Acharai et al., 2023). And while there is evidence to suggest progress in improving access to these services (WHO,2022), significant barriers remain. Specifically, poor knowledge and access to information, sociocultural norms, stigma, shame, lack of social support, lack of infrastructure and security, and GBV continue to create health disparities for women and girls seeking SRH care in Morocco (Acharai et al., 2023; Tirado et al., 2020). Moreover, current research on GBV in Morocco overwhelmingly focuses on sexual or physical violence, resulting in key gaps in our understanding of reproductive violence in this context (Gagliardi, 2018). Accordingly, Morocco is still off=target for achieving its SDG 5 objective of achieving gender equality and empowering all women and girls by 2030.

The number of migrants to Morocco has grown over the last decade. The International Organization for Migration (IOM), for example, estimates that Morocco was home to 102,000 migrants as of 2021 (Acharai et al., 2023). L’Association Marocaine de Planification Familiale (AMPF) is one of the main providers of SRH services in Morocco for both Moroccan and refugee populations. Over the course of their work, AMPF have indicated that RV is near universal among refugee and migrant women, many of whom are hindered from accessing and using their desired method of contraception (Acharai et al., 2023).

Taken together, progress in protecting individual rights to reproductive health hinges on the emergence of systemic shifts that transcend individual, societal, and political domains.

Underpinning these efforts is the need to advance our understanding of how individuals in different social and cultural contexts experience RV and related experiences of RC. Currently, findings in the international literature regarding RV are difficult to generalize as the contexts in which the studies were carried out vary in terms of gender norms, religious influence, legal status of abortion, and socioeconomic status (Grace & Anderson, 2018). Moreover, and despite evidence to support a definition of RV that encompasses multiple potential perpetrators beyond intimate partners, current screening tools tend to focus solely on aspects of reproductive coercion that are perpetrated by intimate partners (e.g., condom sabotage), and do not explicitly account for other acts of reproductive violence (DeJoy, 2019; Grace & Anderson, 2018). The current study thus aimed to develop an understanding of the ways in which RC could be perpetrated by other actors and through less obvious means. Clarity regarding the nature of RV and its associated factors is thus critical to increase the visibility of RV, inform more targeted and effective interventions, and develop valid and reliable assessments of RV that can be used for both screening and monitoring and evaluation purposes.

In light of these gaps, the present studies overall aim is to generate a stakeholder-informed understanding of reproductive violence as it appears in the Moroccan context. Specifically, we sought to develop a context-specific screening tool to ensure that assessments of RC are fit-for-purpose and offer a more reliable assessment of the extent and nature of RV experienced by women (including refugee and migrant women) in Morocco.

## Method

### Study design and procedures

Delphi methods were employed over two rounds of consultation to determine the level of consensus on and stakeholder perceptions of a series of key statements regarding reproductive violence and coercion in the Moroccan context. A Delphi approach was selected given its widespread use in the development of new measures or tools (Barratt & Heale, 2020).

#### Generation of Item Statements

Statements were developed over several steps. First, we conducted a rapid review of the literature to identify the extent, range, and nature of relevant research evidence regarding RV and RC, and to clarify the working definitions of these concepts. Screening tools, questionnaires and surveys related to RC previously used in survey research and clinical practice were also identified. Next, rapid review results and identified tools were analyzed during workshop style focus groups using thematic analysis (Braun & Clarke, 2006). Results were used to develop a conceptual framework, identifying examples of reproductive violence relevant to the Moroccan context that fell under each of the following levels: 1) reproductive coercion perpetrated by intimate partners and family, 2) social norms, and 3) institutional violence.

A set of initial 29 item statements was generated The items were developed in English, translated to French, and back-translated by members of the research team prior to presenting these to the project’s stakeholder advisory group for discussion and expansion. The advisory group consisted of an international group of practitioners, academics, advocates, and health professionals familiar with the fields of GBV and SRH. Formed at the outset of the project, the advisory group functioned as an expert steering committee, meeting regularly throughout the project for support and consultation, lending their expertise to guide methodological and ethical decisions, and providing additional rigor and insight into the interpretation of results. At item development stage, discussions among the advisory group resulted in the generation of four additional statements and several suggested edits to the original 29 statements. A total of 33 final statements were thus included in the first round of the Delphi study (12 statements falling under level 1, 13 under level 2 and 8 under level 3).

#### Participants

Participants for the Delphi study were selected purposively for their expertise and knowledge of SRH and GBV in Morocco, including AMPF health providers, volunteers, NGO and governmental providers, policy makers, in addition to members of the advisory group.

Organizations relevant to SRH and GBV, advocacy groups for experts by lived experience, alongside representatives from different healthcare professions, including relevant ministries of health, policy makers and other knowledge users were also invited. The recruitment period lasted from 3^rd^ October 2023 until 27^th^ March 2024. Potential participants were sent an invitation email with the participant information leaflet in attachment. Those who indicated willingness to take part (*n* = 23) were then sent a consent form and all participants provided written consent. Of these, *n* = 11 participated in the first round of the Delphi. Only those who had participated in the first round could join the second round. A total of *n* = 10 participants completed both rounds. Of the sample who provided data for the first round, *n* = 9 self-identified as female, and *n* = 2 identified as male. Geographic contexts where participants conducted their work on SRH or GBV included North Africa (n=6), East Africa (n=1), South Asia (n=1), North America (n=1), and countries spanning various regions (n=1). One participant did not specify. Roles identified included researchers, health providers, and program coordinators, many with dual roles.

#### Round 1 – Delphi

Participants who completed the consent form were directed to the specialised online platform for Delphi surveys, Welphi (www.welphi.com), and were all invited to complete their scoring on the same date. Participants were then directed to the statements, presented in both English and French, and were asked to rate each statement using a Likert scale, ranging from “strongly disagree” (= 1), “disagree”, “neutral”, “agree”, “strongly agree” (= 5). Participants were further encouraged to comment in support of their selection for each statement to capture their reflections on the face-validity of each statement. Participants were given a two-week window to complete this first survey round. Voting for the first round was closed after two weeks.

#### Round 2 – Delphi

Statements which deemed to have reached clear consensus in Round 1 were removed for Round 2 to avoid survey fatigue. Participants then received a new link to participate in a second round, where the process of voting and commenting on the statements was repeated. This round was also open for a two-week period. In this second round however, participants could view the anonymous votes and comments of the other participants from the previous round and decide whether to revise or maintain their previous scoring.

### Data analysis

Consensus was defined as 75% who indicated ‘strongly agree’ or ‘agree’ for a given item. The 75% cut-off was selected as the conventional cut-off point for Delphi studies (Diamond et al. 2014). Final consensus scores for all 33 statements are presented in results, alongside relevant comments submitted by participants regarding their perceptions of the statements’ face validity and their relevance to the Moroccan context.

### Ethical Considerations

Ethical approvals for data collection relevant to this project were obtained from Comité d’Ethique pour la Recherche Biomédicale d’Oujda Faculté de Médecine et de Pharmacie Université Mohammed Premiere and the Trinity Centre for Global Health Policy Management/Centre for Global Health Research Ethics Committee on October 2^nd^, 2023 (Approval Number: 2384).

## Results

There was a high degree of consensus across most of the statements presented, with 23 statements passing the 75% threshold across ‘strongly agree’ and ‘agree’ votes by Round 2. Five statements deemed to have reached clear consensus in Round 1 were removed for Round 2 to avoid survey fatigue. See Table 1.

**Table 1.** Final consensus scores of all other statements.

| Statement | Consensus Round 1 | Consensus Round 2 |
| --- | --- | --- |
| ***Reproductive violence is present when women are told they will be divorced, or their partner will take a second wife if they become pregnant or if they do not become pregnant.*** | 100% | N/A |
| ***Reproductive violence is present when women are not permitted to attend reproductive health centres to access contraception*** | 100% | N/A |
| ***Reproductive violence is present when a partner refuses to wear a condom with the intention of impregnating the woman.*** | 100% | N/A |
| ***Reproductive violence is present when migrant or refugee women are given contraception without their consent.*** | 90% | N/A |
| ***Reproductive violence is present when a woman is threatened with or fears experiencing physical violence if she does or does not become pregnant*** | 78% | N/A |
| **Reproductive violence is present when women fear marginalization (or exclusion) by their community if they become pregnant or if they don’t.** | 88% | 100% |
| **Reproductive violence is present when women are not permitted to visit a reproductive health center at all or if she is unaccompanied.** | 77% | 90% |
| **Reproductive violence is present when a partner refuses to pull out while using ‘safe period’ as a contraceptive method, with the intention to impregnate the woman.** | 89% | 90% |
| **Reproductive violence is present when a woman fear she will be denied economic resources if she does or does not become pregnant.** | 77% | 90% |
| **Reproductive violence is present when a woman is pressured to continue to have children until she gives birth to a son or to a girl.** | 89% | 90% |
| **Reproductive violence is present where when married women are denied access to contraception due to the belief that contraception means that they are more likely to cheat or have partners other than her husband.** | 77% | 90% |
| **Reproductive violence is present when women are not involved in decision making about contraceptive use or about the form of contraception used.** | 77% | 80% |
| **Reproductive violence is present when a woman fears experiencing psychological abuse (such as exclusion or shaming) if she does or does not become pregnant.** | 89% | 80% |
| **Reproductive violence is present when women’s value within the household is measured by her fertility.** | 78% | 80% |
| **Reproductive violence is present where women are prevented from working or going to school to increase their chances of becoming pregnant.** | 66% | 80% |
| **Reproductive violence is present where contraception is denied due to a belief that contraception impacts on a woman’s fertility.** | 78% | 80% |
| **Reproductive violence is present where when unmarried women are denied access to contraception due to the belief that contraception will increase a woman’s promiscuity and/or ability to have relationship with multiple partners.** | 78% | 80% |
| **Reproductive violence is present where contraception is denied due to a belief that contraception is bad for married women’s reputation.** | 66% | 80% |
| **Reproductive violence is present when migrant or refugee women do not have access to sexual and reproductive health services due to their migrant/refugee status.** | 89% | 80% |
| **Reproductive violence is present when women fear marginalisation (or exclusion) by their family or partner if they become pregnant or if they don’t.** | 80% | 80% |
| **Reproductive violence is present when a health provider pressures a woman to use a particular type of contraception.** | 78% | 80% |
| **Reproductive violence is present when women with disabilities are given contraception without seeking or consenting to it** | 77% | 80% |
| **Reproductive violence is present when women who have been raped/ experienced sexual violence do not have access to emergency contraception (EC).** | 78% | 80% |
| Reproductive violence is present where women need to have at least one child before they can be offered or given access to contraception. | 77% | 70% |
| Reproductive violence is present when a woman's partner/mother-in-law/others tell her to use or not to use contraception. | 78% | 70% |
| Reproductive violence is present where institutions prevent women from returning to work, or to education during or after pregnancy. | 55% | 60% |
| Reproductive violence is present where an existing pregnancy is used to justify the registration of a child marriage. | 33% | 50% |
| Reproductive violence is present when there are no maternity leave policies or social protection measures for pregnant women or women with children, especially in the informal and agricultural sectors. | 44% | 50% |
| If reproductive violence is present, then unmarried women who become pregnant will be forced to marry. | 33% | 40% |
| If reproductive violence is present, then unmarried women who become pregnant will be cut off from their family members. | 33% | 40% |
| Reproductive violence is present in the instance where a person under the legal age of consent (18) is pregnant. | 22% | 30% |
| Reproductive violence is present when women who have been raped/ experienced sexual violence and then become pregnant will be forced by law to marry their partner. | 33% | 30% |
| Reproductive violence is present when it is believed that women with disabilities should not have children. | 33% | 20% |

There were five statements achieving over 75% for the option of ‘strongly agree’ alone in round 1 – and thus not included in round 2. These included, “Reproductive violence is present when women are told they will be divorced, or their partner will take a second wife if they become pregnant or if they do not become pregnant” (91% ‘strongly agree’ in Round 1). Participants elaborated on how this form of RV was relevant to the Moroccan context:

> *The cultural emphasis on lineage and inheritance is significant, the pressure on women to bear children to continue the family name can be immense. When a woman is threatened with divorce…it places her reproductive decisions under the control of her partner’s desires, undermining her autonomy and rights. This…reflects broader societal expectations and norms around marriage, fertility, and women’s roles.*

For another statement, “Reproductive violence is present when women are not permitted to attend reproductive health centres to access contraception”, and which achieved 100% consensus in Round 1, respondents identified how this statement is indicative of RV perpetration influenced by social norms:

> *Unless your family is respectful of you and your decisions (rare if patriarchal and husband decisive on contraception, etc.) accompanied will not work. This is true especially for adolescents. We also have…qual[itative] data about family or partner accompanying [women] to SRH appointments to ensure they did not disclose sexual violence*.

Other statements which reached 100% consensus with a score of 80% for strongly agree in Round 1 related to more direct sexual acts: “Reproductive violence is present when a partner refuses to wear a condom with the intention of impregnating the woman” and ‘’Reproductive violence is present when a partner refuses to pull out while using ‘safe period’ as a contraceptive method, with the intention to impregnate the woman”. For these, participants highlighted social norms within Morocco, and gendered power imbalances more broadly as key factors relevant to these statements:

> *In Morocco, as in many parts of the world, relationships where men prioritize fulfilling their desires regardless of their partner’s thoughts or consent highlight significant issues of power imbalance and lack of respect for women’s autonomy and rights*.

An overlap between RV and other forms of violence was identified within several statements. For example, “Reproductive violence is present when a woman is threatened with or fears experiencing physical violence if she does or does not become pregnant” achieved a 78% consensus in Round 1, where all scores fell under ‘strongly agree’ with participants indicating that threats of physical violence clearly “violate a woman’s reproductive autonomy and deny her the fundamental right to make decisions about her own health and life”. One statement relating to psychological abuse “*Reproductive violence is present when a woman fears experiencing psychological abuse [such as exclusion or shaming] if she does or does not become pregnant*” received a higher consensus score of 89%, but only 33% of participants indicated strong agreement. Participant comments for this statement highlight the variation in perpetrator identity: “people spoke about being ‘deathly afraid’ if their mother, for example, would never speak to them again, so they did what she wishes”. In addition, comments to this statement highlighted several cultural elements deemed to be specific to a Moroccan context:

> *Especially those who cannot get pregnant because culturally it is always the woman who is at fault (for some people there is confusion between virility and fertility) so these women can be victims of psychological and economic violence… because they are afraid of being repudiated*.

Economic violence was also identified separately within the study with the statement “Reproductive violence is present when a woman fears she will be denied economic resources if she does or does not become pregnant’’ reaching 90% consensus, with participants highlighting the complexity of this form of RV regarding sources of perpetration and the interaction with other forms of structural inequalities:

> *In some cases, women may fear losing their jobs or suffering discrimination in the workplace if they become pregnant. At other times, they may fear economic consequences if they choose not to have children, such as the loss of financial support from a partner or family. These economic pressures limit women’s reproductive autonomy and also contribute to perpetuating gender inequalities*.

Participants also endorsed statements which considered intersectional oppression and acknowledged the ways in which women with multiple vulnerabilities, complex needs or intersecting minoritized identities are affected, such as ‘’Reproductive violence is present when migrant or refugee women are given contraception without their consent’’ (90%) and ‘’Reproductive violence is present when women with disabilities are given contraception without seeking or consenting to it’’ (77%).

Acknowledgement of the broad range of actors who perpetrate RV was captured in the statements “Reproductive violence is present when women fear marginalization (or exclusion) by their family or partner if they become pregnant or if they don’t” and “Reproductive violence is present when women fear marginalization (or exclusion) by their community if they become pregnant or if they don’t”, which reached 90% and 88% consensus, respectively.

Several statements which reached consensus focused specifically on social norms regarding gender and reproduction. These included the statement “Reproductive violence is present when a woman is pressured to continue to have children until she gives birth to a son or to a daughter”, which reached 90% consensus, with several participants indicating in comments that within a Moroccan context this is “’More precisely, until she gives birth to a boy”. Participants also elaborated on the social and gender norms influencing this form of RV:

Multiple patriarchal “reasons” justify this reproductive violence: the girl risks dishonour, the perpetuation of the lineage, the boy as old-age insurance, the integrity of the inheritance… This is changing in the sense that girls are increasingly perceived as being more tender and more inclined to take care of their elderly parents.

Also reaching consensus were statements capturing beliefs and attitudes regarding female fertility. These included “Reproductive violence is present when a woman’s value within the household is measured by her fertility” (80% consensus), which participants noted was “a subtle form of violence”, which “can create a huge amount of pressure on women to conform to restrictive social expectations of reproduction, and to define themselves primarily by their ability to have children”. Participants also noted that within Morocco this was a “common practice” due to:

> *Traditional values [that] often emphasize the importance of motherhood, women may face significant societal and familial pressure to fulfill these roles. This scenario clearly constitutes reproductive violence by enforcing a narrow, fertility-based valuation of women, thereby impacting their physical and psychological health and well-being*.

The impact of these beliefs, coupled with what participants identified as a ‘lack of comprehensive sexual education (CSE)’ can be seen in the statement “Reproductive violence is present where contraception is denied due to a belief that contraception impacts on a woman’s fertility”; (80% consensus). However, another participant disagreed that lack of education was a causal factor, reporting that “some women seeking contraception from the (AMPF) without their husbands’ knowledge, highlights a significant issue around reproductive autonomy and gender dynamics despite the accessibility to information and the digitalization”.

These gender dynamics are evident in cultural attitudes regarding sexuality, particularly those which lead to the denial or control expression of female sexuality, and are captured in the statements “Reproductive violence is present where when married women are denied access to contraception due to the belief that contraception means that they are more likely to cheat or have partners other than her husband” (90% consensus), and “Reproductive violence is present where when unmarried women are denied access to contraception due to the belief that contraception will increase a woman’s promiscuity and/or ability to have relationship with multiple partners” (80% consensus).

One participant discussed these statements in the context of Moroccan cultural values regarding extra-marital sexuality:

> *Access to contraception is a SRH right, even if the Sharia and the Penal Code prohibit any non-marital sexuality. Public SRH services must ensure access to contraception for all women of reproductive age (even if clandestinely). And even if it is clandestine, public authorities must include in their statistics the number of unmarried women who use contraception. These authorities must not officially limit themselves to leaving the private sector the responsibility of providing contraceptives to unmarried women*.

However, participants highlighted that it was not only intimate partners who enacted RV due to beliefs regarding female sexuality. They indicated that “in some cases, this practice exists in healthcare settings, stigmatizing unmarried women for extramarital relationships”. This comment aligns with the statement “Reproductive Violence is present when a health provider pressures a woman to use a particular type of contraception” (80% consensus), which participants identified as ‘institutional reproductive violence’. Participants elaborated on this, stating that even if the pressure was for medical reasons, “We should not exert pressure but explain and help the person to make an informed choice and leave them free to choose.’’

There were two statements which did not reach the threshold in the first round but did in the second round. These include “Reproductive violence is present where contraception is denied due to a belief that contraception is bad for married women’s reputation”, which achieved a score of 66% in Round 1, which rose to 80% in Round 2. One participant explained their choice not to endorse this statement, stating “The definition of reproductive coercion typically requires intent to make her pregnant against her wishes. If a husband prevents her from using a [family planning] method for other reasons (like concerns about her reputation), we have termed this contraceptive-related coercive control” while another participant who strongly agreed stated that it is “not about the reason or the belief but the ‘denial’ and whether or not the belief is used to obstruct someone’s SRHR”. The statement “Reproductive violence is present where women are prevented from working or going to school to increase their chances of becoming pregnant”, which similarly rose from 66% to 80% across both rounds. Participants expressed that reservation with this statement were due to a perceived conflation of RV and gender inequality.

Two statements which reached consensus in Round 1 subsequently failed to pass the threshold in the Round 2. These include “Reproductive violence is present where women need to have at least one child before they can be offered or given access to contraception”, which fell from 77% to 70% in Round 2 and “Reproductive violence is present when a woman’s partner/mother-in-law/others tell her to use or not to use contraception”, which dropped from 78% to 70%. Issues identified with both statements include a lack of clarity around the coercive or ‘demanding’ element of the statement and that these statements should make explicit the power dynamics in each situation.

Several statements did not meet the threshold in either round, with comments indicating that participants felt the situations described would need to be examined on a case-by-case basis to determine whether RV was present. Statements included “If reproductive violence is present, then unmarried women who become pregnant will be forced to marry” (40%). One participant explained their hesitancy to endorse this statement, saying:

> *This is a tricky one - I believe forced marriage is GBV but because it is due to pregnancy whether that then makes it RV, I’m not sure. I conceptualize RV more as related to the pregnancy outcome in this scenario. So not the forced marriage, but RV in the case of threats e.g. ‘if you continue this pregnancy (or if you ever become pregnant) you will have to marry this person.’ (i.e. instilling fear to prevent pregnancy)*.

Other statements failing to meet the threshold included “If reproductive violence is present, then unmarried women who become pregnant will be cut off from their family members” (40%), which participants noted would not ‘’automatically be deemed RV, it depends on the case’’ and “Reproductive violence is present where an existing pregnancy is used to justify the registration of a child marriage” (50%), which one participant queried, saying ‘If the person ends up having a miscarriage before the wedding, or they find an unsafe abortion provider and are no longer pregnant, can they get out of the marriage arrangement?’.

## Discussion

This Delphi study aimed to advance our understanding of reproductive violence in the Moroccan context through determining the level of consensus and analysing participant comments for 33 item statements regarding reproductive violence and coercion.

Several of the statements reaching consensus map onto existing questions included in the 9-item Reproductive Coercion Scale (RCS) (Miller et al., 2010), a commonly used scale to screen for RC experiences. Specifically, high consensus was found with comparable RCS items 1-6, which include: ‘(Your Partner) Told you not to use any birth control (like the pill, shot, ring, etc.)’, ‘Said he would leave you if you didn’t get pregnant’, ‘Told you he would have a baby with someone else if you didn’t get pregnant’, ‘Taken your birth control (like pills) away from you or kept you from going to the clinic to get birth control’, ‘Made you have sex without a condom so you would get pregnant’, and ‘Hurt you physically because you did not agree to get pregnant’.

The final set of consensus statements highlight important social and cultural norms and ideas about gender roles which influence the perpetration of RC not currently captured within common screening measures. This included cultural emphasis on lineage, societal expectations for women to assume the role of wife and mother, and patriarchal norms which privilege the desire of men over the reproductive or sexual autonomy of women. This is reflective of findings reported by Ouahid et al. (2023) where access to contraceptives for women in Morocco was found to be restricted by gender norms; for young or unmarried women to access these services would be akin to ‘societal suicide’. This restriction on reproductive autonomy is not enforced exclusively by intimate partners, but also by family and communities, which the current study also found marginalize or exclude women who transgress these gender norms. Previous studies based in Morocco have identified similar experiences, with Ouahid and colleagues (2023) also finding that peer and community pressures influence the decisions of married women regarding contraceptive access, discouraging them from using contraception until they give birth to a male child. Indeed, RC occurs due to an interplay between individual, relationship, community, and societal factors, with abusive behaviors arising from and being reinforced by society and culture (Graham, et al., 2023). Our findings illustrate the importance of considering not only individual behaviours but also wider societal factors that influence reproductive autonomy such as gender norms, policies, and cultural beliefs, and integrating these factors into both screening and intervention tools.

Our findings also show considerable overlap between RC and RV and other forms of gender-based violence and abuse, including financial, psychological and physical. This is illustrated by participants highlighting that not adhering to gender role norms or complying with expectations of male partners or family regarding reproduction can place women at risk of psychological abuse, including exclusion or shaming. Participants also identified that removal of resources and finances can be used to enable reproductive coercion. This aligns with other research which demonstrates a strong overlap between intimate partner violence (IPV) and RC (Grace & Anderson, 2018), however more research is needed to elucidate the exact nature of how reproductive coercion and other forms of GBV interact and co-occur. This is currently poorly understood, impeding identification of causal mechanisms and risk factors for reproductive coercion and hindering the development of effective and targeted interventions and supports (Muñoz et al., 2023).

Finally, the current study highlighted how RC can be perpetrated against specific vulnerable populations. Delphi participants endorsed several statements indicating that RV is present when migrant or refugee women, or women with disabilities are given contraception without seeking or consenting to it. Understanding how migrant women in particular experience RC is important within the Moroccan context, as a recent study conducted with 151 female migrants in Rabat highlighted that 87.4% of respondents had experienced SGBV during their lifetime, 58.6% declared having been threatened with physical force. The study also found that despite community-based health services launched by the Moroccan Association of Family Planning, there is an underutilization of healthcare services by the migrant population (Acharai et al., 2023). This points to the need for research which explicitly incorporates an intersectional approach to explore the ways in which multiple inequalities intersect to create specific vulnerabilities to gender-based violence (Humbert et al., 2024). The literature is clear that membership in certain minoritized groups is associated with higher exposure to gender-based violence, including LGBT communities, ethnic and religious minority groups and migrants and people with disabilities (Messinger, 2011; Roudsari et al., 2009; Voolma, 2018). Failure to integrate the lived experiences of those with intersecting identities in research and intervention development results in a lack of acknowledgement of the complexities of social and cultural forces underpinning reproductive coercion which are played out at the interpersonal level. More work is needed to develop contextually tailored and inclusive interventions, and training which empowers providers to interact with and respond to service users in a manner that is culturally and contextually relevant and appropriate (Grace & Miller, 2023). Overall, our results suggest that, in Morocco, RC and RV are intimately linked to other intersecting forms of oppression and contribute to perpetuating gender inequalities.

### Limitations and Future Considerations

This study generated data regarding stakeholders’ understandings of reproductive violence as it is experienced in a Moroccan context. However, several limitations to the current study are noted. First, the final sample size was relatively small. While there is no standard sample size for Delphi studies, the typical range is between 10-100 panelists, with considerations such as complexity of the topic, heterogeneity of the expert panel, and availability of resources impacting numbers. While the current study sought to recruit stakeholders with specific expertise relevant to a particular context, increased sample size would have enhanced the generalizability and reliability of our findings. The sample also included a disproportionate number of respondents who self-identified as female, while the self-selecting nature of the panel meant that those with a particular interest in the topic were more likely to respond to invitations to take part. Further studies could seek to expand the number of participants overall and take measures to ensure a greater gender balance. Additionally, due to time and resource constraints a pilot study was not conducted, which may have enhanced the clarity and accessibility of the final statements. This would be particularly relevant when presenting statements across two languages to ensure there are no translation issues. Finally, future studies should seek to ascertain the validity of a scale which incorporates all 33 item statements through subjecting it to additional psychometric testing as a next step in the development of a screening tool.

## Conclusion

This study provides important insights into how reproductive coercion and violence are experienced in the Moroccan context. While findings indicate several elements of reproductive coercion that are consistent with current screening tools and research findings, additional statements capture the specific socio-political landscape of the country and how this shapes the forms of RV that are experienced by women and girls. Further examination of any differences in experiences of RV within refugee and migrant communities and across both urban and rural divides in Morocco may be a useful priority for future research, to ensure contextualized screening tools for RV within AMPF, enabling the provision of interventions and support to all affected clients.

## Data Availability

Data will be submitted to TARA. TARA is an institutional repository designed to store, catalogue, index, distribute, and preserve the research outputs of Trinity College Dublin. It includes the full text of peer-reviewed research publications, conference papers, books and book chapters, entire journals, electronic theses, working papers and technical reports and images. Unless specifically restricted, material in TARA is Open Access i.e., fully accessible via Google, Google Scholar etc. and exposed for harvesting by search engines, and web-based bots and harvesters.

## Notes

**Disclosure statement:** The authors report no conflict of interest.

### Competing Interest Statement

The authors have declared no competing interest.

### Funding Statement

Yes

### Author Declarations

Ethical approvals for data collection relevant to this project were obtained from Comité d’Ethique pour la Recherche Biomédicale d’Oujda Faculté de Médecine et de Pharmacie Université Mohammed Premiere and the Trinity Centre for Global Health Policy Management/Centre for Global Health Research Ethics Committee on October 2nd, 2023 (Approval Number: 2384).

